# Perspectives and use of telemedicine by doctors in India: A cross-sectional study

**DOI:** 10.1101/2022.09.30.22280571

**Authors:** Vikranth H. Nagaraja, Biswanath Ghosh Dastidar, Shailesh Suri, Anant R. Jani

**Affiliations:** Department of Engineering Science, Institute of Biomedical Engineering, University of Oxford, Oxford OX1 3PJ, UK; Oxford India Centre for Sustainable Development, University of Oxford, Oxford OX2 6HD, UK; GD Institute for Fertility Research, Kolkata – 700025, India; Cambridge University Hospitals, Cambridge, CB20QQ, UK; Heidelberg Institute for Global Health, Heidelberg, Germany

**Keywords:** COVID-19, Digital health, Healthcare delivery, Healthcare policy, India, Internet, Telemedicine

## Abstract

**Objectives:** India has committed to formulating a roadmap for realising a resilient health system, with digital health being an important element of this. Following the successful implementation of a free telemedicine service, eSanjeevani, India published the Telemedicine Practice Guidelines in 2020 to further scale telemedicine use in India. The main objective of the current study was to understand the perspective and use of telemedicine by medical doctors in India after the release of its telemedicine policy.

**Methods:** Data were acquired through an anonymous, cross-sectional, internet-based survey of medical doctors (n = 444) at a pan-India level. Replies were subjected to statistical analysis.

**Results:** Telemedicine was used for various non-mutually exclusive reasons, with the top two reasons being live audio or video consultations (60.4%) and online payments (19.1%) and smartphones were the most frequently used device type (60.6%). The telemedicine benefit that the greatest proportion of respondents (93%) recognised was its potential to reduce COVID-19 infection risk for healthcare professionals. Interestingly, nearly 45% of respondents felt that limited and fragmented insurance coverage was an important limitation to the practice of telemedicine in India and 49% believed reduced patient fees for teleconsultations could help incentivise telemedicine use.

**Conclusions:** This study helps to appraise the use of telemedicine in India after the publication of telemedicine guidelines in 2020. Furthermore, the findings can inform the development of telemedicine platforms, policies and incentives to improve the design and implementation of effective telemedicine in India.

**Public Interest Summary:** India has committed to formulating a roadmap for realising a resilient health system, with digital health being an important element of this. In 2020, India published its Telemedicine Practice Guidelines to scale telemedicine use in India. The main objective of the current study was to survey medical doctors in India to understand their perspectives on and use of telemedicine after the release of India’s telemedicine policy. Our findings revealed that the top two reasons doctors used telemedicine were for live audio or video consultations and online payments. Interestingly, a large proportion of respondents felt that limited and fragmented insurance coverage was an important limitation to the practice of telemedicine in India. This study helps to appraise the use of telemedicine in India after the publication of its telemedicine guidelines and can inform the development of telemedicine platforms, policies and incentives to improve the design and implementation of telemedicine in India.

## INTRODUCTION

In 2018, India achieved the WHO-recommended doctor-to-population ratio of 1:1000 – an impressive feat for a country with a population as large as India’s.^1,2^ While this is an important step, significant problems persist in India including substantial unmet need, poor health outcomes, as well as large and increasing health inequalities across India.^3,4,5^ India also accounts for one of the highest out-of-pocket expenditures worldwide, which can be catastrophic or impoverishing and disproportionately burdens those from low-income backgrounds.^6^ Ambade et al.^7^ analysed the components of health expenditure (particularly out-of-pocket expenditure) and their relative contribution to the economic burden of diseases in India, with travel, lodging, and food accounting for a significant proportion of non-medical costs linked to accessing care. These negative trends have been exacerbated by the COVID-19 pandemic and its associated lockdowns.^8^ In a country as large, heterogeneous, and complex as India, given the current healthcare training infrastructure, it may be challenging to scale up the number of healthcare professionals fast enough to meet increasing healthcare need and demand, especially in rural and isolated regions.^9^

Digital health (which includes technologies such as telemedicine, mobile phones and associated applications, Internet of Things [IoT], wearable devices and sensors, robotics, virtual and augmented reality, and AI and genomics) has been framed as a strategic priority for healthcare systems across the world because of its potential to augment and transform existing care pathways and care delivery models.^10^ In 2005, the World Health Assembly urged member states to develop long-term strategic plans for digital health.^11^ Due to the growing influence that different aspects of digital technology access and literacy might have on health and well-being outcomes, Kickbusch et al.^12^ note that the digital ecosystem can be considered an increasingly important determinant of health.

At a population and infrastructural level, India is well-positioned to adopt and transition to a healthcare system underpinned by digital health. India is reported as one of the fastest-growing digital consumer markets, with over 0.5 billion internet subscribers using more than 8GB/month, and it has some of the lowest data costs in the world.^13^ Pew Research Center reports that mobile phone ownership, particularly smartphone ownership and internet usage, has continually grown in the recent decade in emerging economies like India.^14^ Furthermore, the Government BharatNet program^15^ intends to extend the fibre network to all Indian villages by 2025, and it is predicted^16^ that, consequently, India will have one billion smartphone users by 2026.

One of the earliest formal forays into digital health in India came in 2019 when the state of Andhra Pradesh piloted *eSanjeevani*, a free telemedicine initiative. Though there have been similar initiatives to increase the use of telemedicine in several resource poor settings, policy and regulatory challenges have impeded their widespread adoption.^17^ In India, for example, particular impediments included a lack of legal and administrative frameworks, a lack of clarity on implementing telemedicine modalities for healthcare service delivery, as well as inertia from healthcare service providers to adopt recent developments in information and communication technologies.^18^ This resulted in poor and heterogeneous adoption and integration of telemedicine services across India. The COVID-19 pandemic and its associated lockdowns, however, provided an opportunity to showcase the role telehealth can play in strengthening often neglected and under-funded primary care services in India.^19^ *eSanjeevani* was adopted at a national level as the pillar for India’s National Telemedicine Service in 2020 by the Ministry of Health and Family Welfare (MoHFW),^20^ which was complemented in 2020 with the flagship National Digital Health Mission initiative, which aims to create a digital health ecosystem for the country to support universal healthcare and improve health outcomes.^21,22^ On March 25, 2020, the Telemedicine Practice Guidelines^23^ were released by the MoHFW (Government of India) and the Medical Council of India for Registered Medical Practitioners (RMPs). These guidelines offer clear, practical advice to RMPs about using technological advancements in medical practice and serve as an essential first step in organising telemedicine services nationwide. Notably, the scope, definitions, norms, protocols, and framework to implement telemedicine services were outlined in these guidelines. They also clarified the roles and responsibilities of RMPs, patients, health workers, and telemedicine platforms while administering/receiving telemedicine-based care and services. Since its launch, there have been over thirty million teleconsultations through *eSanjeevani*. It is important to note, though, that this is a tiny fraction relative to the total need and demand for healthcare services in India.^20^ Furthermore, serious questions exist about the on-the-ground realities surrounding the use and utility of digital health modalities like telemedicine.

To improve our understanding of the role that telemedicine can practically play in India’s healthcare systems, we surveyed practising doctors across India to gauge their telemedicine practices; enablers, barriers and preferences related to the use of telemedicine; and their perspectives on its role in India’s healthcare system after the publication of India’s Telemedicine guidelines.

## METHODS

### Study design

As this study focussed on medical doctors, patients or the general public were not involved in the study design. The details of the study design were as follows:

- Study design: Descriptive study (Questionnaire-based survey)
- Time perspective: Cross-sectional
- Target population sampling: Non-probability sampling (a combination of Convenience sampling and Snowball sampling)
- Study exclusion criteria were:
  - medical doctors who were not aware of telemedicine;
  - doctors not using telemedicine in their clinical practice (either prior to or at the time of the survey); and
  - those who did not complete the entire survey.

The methods and study findings are reported in adherence to the CHecklist for Reporting Results of Internet E-Surveys (CHERRIES)^24^ as well as recommended guidelines for conducting and reporting survey research.^25^

### Ethical approval

This study aimed to survey medical doctors anonymously via a questionnaire to review their current use of telemedicine. This study did not involve patient participation, material or seek identifiable, personal and/or confidential information from the survey respondent. Hence, no ethical approval was required for this study according to the Declaration of Helsinki of the World Medical Association. In addition, upon enquiry from the local ethics committee at the University of Oxford, this study was deemed a ‘service review’ (confirmation available upon request).

The General information on the online survey landing page and the Privacy notice on the second survey page to potential participants explained the reasons for the survey, the details of the investigators, and the fact that participants’ anonymised responses may be published. In addition, the Privacy notice detailed: how the authors collected data from the survey responses; which data was going to be stored; where it was going to be stored and for how long; and how the study team was going to use the data in their analyses. The survey started once the participant agreed with the stated terms and conditions. Informed consent to participate and publication of results was implied through their completion of the study questionnaire.

### Survey design

An open, online, self-administered survey was used to collect quantitative and qualitative data from participants anonymously with no geographic restrictions for respondents (within India) and with no specified timeframe (i.e., the time people needed to fill in a questionnaire completely). Among other reasons, the survey was made anonymous to reduce the risk of desirability bias.^26^

Two of the study authors (VHN and ARJ) conducted a literature scan in PubMed for articles published between 2010–2019 to identify key themes in the area of telemedicine policy and practice. Between them, they developed a consensus around the key themes and designed a draft questionnaire in English based on this exercise. To assess the usability and technical functionality of the survey, a draft version of the survey was tested with five doctors from the authors’ personal and professional networks in India. Minor changes to the survey wording and content were made based on their feedback before finally fielding the questionnaire.

The final survey was designed to take 15–20 minutes to complete and was presented across ten pages (with a range of 1–11 items/questions per page) bookended by the General information/Privacy notice initially and the final (‘Thank you’) page acknowledging survey completion. The survey items—with their order not randomised or modified for different respondents—were categorised into three parts. The first part included items regarding the demographic information of participants. The second part included screening questions pertinent to the COVID-19 pandemic and telemedicine awareness. The third part aimed to elicit information on telemedicine practices; enablers, barriers and preferences related to the use of telemedicine; and respondent perspectives on its role in India’s healthcare system.

The survey consisted of standardised and validated question formats such as (five-point) Likert scales and multiple-choice questions. The respondent had the option to select more than one choice for several multiple-choice questions; for these ‘multi-answer’ questions, percentage values are provided of respondents who selected each answer option (e.g., 100% would represent that all this question’s respondents chose that option). Further, two free-text questions were also included to understand the main challenges with telemedicine that the survey respondents faced at the time of the survey and to seek additional feedback relevant to this study. All questions except those requiring free-text inputs were mandatory to proceed further. Response validation, adaptive questioning, and/or non-response options were used on all questions, where appropriate. Survey progress was displayed on the top of each page. Participants did not have a completeness check/review option at the end of the survey. However, a ‘Finish later’ option was available on every survey page. A copy of the final survey is provided in the supplementary materials (Supplementary Material 1).

### Sampling strategies and target population

A non-probability sampling approach was used for survey dissemination at a pan-India level by contacting the Indian Medical Association, medical colleges in India, the study authors’ personal and professional networks in India, and relevant mailing lists at the University of Oxford and social media. A copy of the advertisement and social media posts containing the exact wording and survey URL is provided in the supplementary materials (Supplementary Material 2).

Participation was voluntary and participants were informed before starting the survey that all data collected was non-identifiable and would be used solely for research purposes. The participants were not reimbursed for their participation. A compulsory selection box consenting to participation was included at the beginning of the survey, ensuring a 100% consent rate. Out of all the study respondents who undertook the study, 139 were automatically screened out based on their responses to the screening questions. After agreeing to participate on Page 2 of the survey, 86 respondents dropped out at various stages (i.e., Pages 3–9) for unknown reasons. In total, 444 medical doctors in India completed the entire survey, a completion rate of 83.8%. Since the survey respondents were not tracked, it is not possible to report suggested information on unique site visitors, view rate, and/or participation rate.^24^

### Data collection

The questionnaire-based survey was developed, hosted, and conducted online on the University of Oxford approved survey tool, *JISC Online Surveys* (https://www.onlinesurveys.ac.uk/) with standard security and confidentiality measures (https://www.onlinesurveys.ac.uk/help-support/online-surveys-security/). This protocol adhered to the General Data Protection Regulation (GDPR) and related UK data protection legislation. Online surveys were designed to protect respondent anonymity. The Online Surveys tool did not use cookies for survey completion and external tracking software such as Google Analytics was not supported on the platform. Additionally, no information about respondents’ Internet Protocol (IP) addresses was collected, thereby making it impossible to gain information on duplicate entries from the same user.

The survey was live to accept responses between 09/12/2020 and 28/02/2021. The automated database for capturing responses was password-protected and stored on secure JISC Online Surveys and an institutionally-issued personal computer of one of the authors (VHN). The survey responses corresponding only to fully-completed questionnaires were exported from the survey tool as a comma-separated values (.CSV) file format for further analysis offline.

### Data/Statistical analysis

All quantitative data analysis was conducted using Microsoft Excel 2016 and MATLAB R2020b (Mathworks, Natick, MA, USA) software. Descriptive statistics (mean, standard deviation, range, frequency count, proportion, and percentage) were used to summarise the survey data.

The responses to Likert-scale questions were aggregated into three categories: ‘Strongly Agree or Agree’; ‘Neither Agree/Disagree’; and ‘Disagree or Strongly Disagree’, wherever applicable. Further, Pareto histograms were used for data visualisation, wherever relevant; however, it should be noted that the questions’ options were not provided in this exact order during the survey (refer to the questionnaire provided in the Supplementary Material for the original sequence of questions).

## RESULTS

### Demographics and respondent characteristics

Of 444 registered medical practitioners who responded to the survey, 91.9% (n = 408) either had been or were, at the time of filling out the survey, on the COVID-19 frontline in India. 59.5% (n = 264) of respondents were men, 38.3% (n = 170) were women; 0.7% (n = 3) identified as “other” and 1.6% (n = 7) preferred not to disclose gender information. The mean ± SD age of respondents was 30.9 ± 9.7 years, with an age range of 20–77 years.

Over two-thirds (68.9%) of the respondents held a ‘Bachelor of Medicine, Bachelor of Surgery (MBBS)’, which is the primary medical degree awarded in the Indian medical education system (accredited by the National Medical Commission [https://www.nmc.org.in/]). Approximately one-third (33.6%) of respondents held either a further diploma or postgraduate medical degree (Supplementary Figure 1).

A majority of respondents were general practitioners (70.9%), with over half the respondents (53.2%) having under five years of clinical experience; while only 4.7% of respondents were active on the COVID frontline were qualified in a medical sub-speciality (Supplementary Figure 2). Less than a quarter of the respondents (21.6%) had ≥15 years of clinical experience (Supplementary Figure 3). Respondents were roughly evenly divided between public (59.7%) and non-public healthcare providers (51.6%) (Supplementary Figure 4). Approximately 81% of respondents were involved in medical practice in nodal urban centres (primarily Mumbai, Navi Mumbai, Kolkata, Pune, Bangalore, and Hyderabad), while the remainder were engaged in practice in rural, semi-urban or community-based practice (Supplementary Figure 5).

### Current telemedicine usage

86.7% (n = 385) of respondents felt they knew the field of telemedicine very well, while 2.5% (n = 11) disagreed or strongly disagreed that they knew the field well.

Respondents had used telemedicine for a variety of different, non-mutually exclusive, purposes including: live audio or video consultations (60.4% [n = 268]); online payments (19.1% [n = 85]); remote self-monitoring (16.7% [n = 74]); home visits by paramedics (16.4% [n = 73]); remote patient monitoring (14.9% [n = 66]); online pharmacy prescriptions (10.1% [n = 45]); store-and-forward technology (9% [n = 40]); and scheduling/changing appointments (8.3% [n = 37]) (Figure 1).

To communicate with patients, respondents reported using a variety of device types and modalities of communication (Figure 2). The majority of respondents reported using smartphones (60.6% [n = 269]) (Figure 2a), while 58.1% (n = 258) of respondents used live audio or video consultations via digital platforms as their primary modality of communication (Figure 2b).

Respondents were asked to estimate the proportion of in-person consultations replaced by virtual consultations in their clinical practice. Although the majority (54.2% [n = 239]) of respondents estimated that less than 25% of their consultations switched from in-person to virtual consultations, this still left a significant number of respondents (45.8% [n = 202]) who estimated that more than a quarter of in-person consultations had been replaced with virtual consultations (Figure 3).

### Respondents’ opinions on Telemedicine practice and policy in India

Respondents were asked to indicate their level of agreement on the different benefits of telemedicine in India (Figure 4). The benefit that the greatest proportion of respondents (93%) strongly agreed or agreed with was the potential for the use of telemedicine to reduce COVID-19 infection risk for healthcare professionals. Interestingly, the benefit that the lowest proportion of respondents (74%) strongly agreed or agreed with—and which the greatest proportion of respondents disagreed or strongly disagreed with—was the ‘potential for telemedicine to deliver better quality of patient care’.

Respondents were also asked to indicate their opinion on different limitations of telemedicine practice in India (Figure 5). Most respondents selected the option of ‘Limited & fragmented insurance coverage of telemedicine,’ with nearly 45% of respondents believing this was an important limitation to the practice of telemedicine in India. The option that was least selected by the respondents was ‘Revise scheduling processes’, with only about 5% of respondents believing this was an important limitation.

Respondents were asked whether they were aware of India’s Telemedicine Practice Guidelines.^23^ Respondents who chose ‘Yes’ (376 of 444) were then asked for their level of agreement on whether these guidelines strengthened telemedicine practice and provided strong medicolegal guidance (Figure 6). Approximately 80% of respondents strongly agreed or agreed that the guidelines strengthened telemedicine practice and provided strong medicolegal guidance.

Respondents were also asked their opinion on policy changes that could further incentivise the use of telemedicine in India (Figure 7). The policy change that the greatest number of respondents (48.7% [n = 206]) believed could help to incentivise telemedicine use in India was reduced fees for teleconsultations for patients. The policy change that the least number of respondents (20.1% [n = 85]) believed could help incentivise telemedicine use was additional incentives commensurate with reduced carbon footprint by the patient.

## CONCLUSIONS

Safe and effective implementation of telemedicine requires an understanding of strategic, operational, and tactical considerations as well as an understanding of on-the-ground realities facing front-line practitioners and patients.^19^ Scott Kruse et al.^17^ highlighted that the top barriers to telemedicine adoption are technology-specific (e.g., technology barriers and lack of computer literacy) and could be overcome through training, change-management techniques, and alternating delivery by telemedicine and personal patient-to-provider interaction, the latter of which is also suggested as a plausible way forward in India.^27^

This study surveyed over 400 medical doctors in India to assess recent trends in telemedicine policy, adoption, practices, and their perspectives after the first wave of the COVID-19 pandemic in India and the introduction of India’s Telemedicine guidelines in 2020.

### Who is using telemedicine in India and where?

Our findings are indicative of the demographics of the COVID-19 response and the prevalence of telemedicine use in India at present. A proportionately greater number of younger doctors were involved in the survey, which, while being mindful of potential sampling bias, could indicate that telemedicine-driven medical practice is higher among younger doctors. Presumably, this segment of the medical workforce is more tech-savvy and has adopted the use of portable tele-devices (smartphones, tablets, etc.) into their regular practice to a greater extent than their more experienced colleagues.

Over 80% of our respondents were involved in medical practice based in urban centres, which confirms previous reports of the urban-rural divide in access to healthcare in India. Interestingly our respondents were nearly evenly distributed between the government and private healthcare sectors, which indicates that telemedicine is a modality of intervention that can cross boundaries between different healthcare providers and provide a common mechanism to bridge the gap between public and private healthcare delivery, especially if a universal patient identifier can be used to capture information on healthcare utilisation and outcomes consistently.

### How is telemedicine being used in India?

Our study revealed that telemedicine-augmented clinical practice was widely used in India (n = 385 [86.7%]) after the first wave of COVID-19. Approximately 45% of all respondents reported that between a quarter and three-quarters of previous in-person appointments had been replaced by virtual consultations, though we cannot be sure if this was before or after the first wave of the COVID-19 pandemic in India.

A variety of technologies were used by doctors for their telemedicine consultations, with approximately 60% of respondents reporting using an Android and/or Apple smartphone and a similar percentage of respondents reporting using audio or video consultations delivered through a digital platform. It is suggested that the number of smartphone users in India will rise from 750 million to one billion by 2026, with the highest growth rates in rural markets.^16^ Given this trend, there is likely significant potential to leverage the use of digital modalities of telemedicine delivery through smartphones in the coming years in India.

### Doctor’s thoughts on the telemedicine policy and practice now and in the future

The first COVID-19 case in India was reported in Kerala on January 29, 2020; the country commenced a 21-day lockdown on March 24, 2020. During this lockdown, most services deemed to be non-essential were suspended, making it extremely challenging to access primary and routine healthcare services. The release of telemedicine guidelines around this period that, among other provisions, provided legal protection to all stakeholders proved to be timely. Over 80% of survey respondents ‘Strongly agreed’ or ‘Agreed’ that these guidelines strengthened telemedicine practice and provided a strong medicolegal framework. The policy changes the greatest number of respondents felt could help incentivise telemedicine use were reduced fees for teleconsultations for patients, followed by indirect benefits for the patients and shorter wait times for scheduling the next appointment.

The surveyed doctors felt the most significant benefit of telemedicine was reducing COVID-19 infection risk for healthcare professionals and perhaps breaking the chain of transmission. The respondents also generally ‘Agreed’ or ‘Strongly agreed’ that telemedicine offered many benefits for all stakeholders involved and helped strengthen capacity. The largest proportion of respondents felt that the biggest limitation in the use of telemedicine was limited and fragmented insurance coverage, which is consistent with our finding that the policy change respondents felt could increase the use of teleconsultations was reduced patient fees.

### Study limitations

Several limitations are acknowledged that reduce generalisability and ready adoption of findings from this study. Firstly, our questionnaire’s psychometric properties (validity and reliability) have not been tested. Second are drawbacks that are common across internet-based surveys, including problems of coverage, lack of suitable sample frameworks, and non-response.^24^ There is also a risk bias being introduced due to (i) increasing the chances of doctors using telemedicine being more likely to be involved in the study and respond to the survey because it is an internet-based survey; (ii) anonymised survey response as well as the choice of sampling strategy; (iii) the non-representative nature of the internet population, and (iv) the self-selection of participants (volunteer effect).^24^ This survey was carried out between the first wave and significantly stronger second wave of the COVID-19 pandemic in India. Given the significant negative impact of the second wave of the COVID-19 pandemic in India, the responses from this survey, particularly on the use of telemedicine, may have been much different after the second wave. The authors hypothesise that usage levels and acceptance may have increased after the second wave, which merits a future study.

### Recommendations for future work

Regardless of the widespread adoption of telemedicine since the pandemic, there are still several challenges that must be overcome to firstly ensure it delivers value and secondly to identify and address the factors that affect the adoption and scaling of the most valuable aspects of telemedicine-augmented care pathways in India.

At the patient-clinician interface, Gudi et al.^19^ note that the mechanisms to evaluate the impact of telemedicine and measure outcomes and acceptability among end-users have not been addressed comprehensively. A survey like the one presented in this study with increased sample size and incorporation of random sampling could help to account for different demographics in India (and perhaps, other South-east Asian countries) to identify and further explore the needs, preferences, and context-specific factors that can influence the use of telemedicine by doctors and other healthcare professionals. Furthermore, a patient-facing survey can help identify the most important factors for patients and identify inequalities in access and use of telemedicine across different patient demographics. It is also important to account for the context-specific factors affecting care-seeking behaviour in India – namely, that approximately 80% of India’s population depends on non-allopathic medicine.^28^ Given this, it is vital to explore the role telemedicine might play in delivering non-allopathic care.

At an organisational level, there is a need to understand how best to design and implement suitable training, enhanced documentation, communication, and observation of information governance guidelines linked to telemedicine use. Furthermore, the success of telehealth could be undermined if strict privacy and security risks are not addressed.^29^ Since telemedicine is an evolving field, especially in a developing country like India, training of medical professionals, clear guidelines, and quality and affordable internet-related infrastructure will facilitate the acceptability of telemedicine in Indian.

At a population and system level, it is important to explore instances of variation, both warranted and unwarranted, in access and outcomes linked to telemedicine use. Identifying warranted variation can help to identify best practices that can be scaled across India and to other care pathways, while identifying unwarranted variation can help to identify health inequalities and inequities. When exploring variation, it is vital to ensure that a comprehensive view is taken of the system at micro-, meso- and macro-levels because various factors can impact the use of and outcomes delivered through telemedicine. For example, at the micro-level, digital literacy and access to training for both healthcare professionals and patients can impact the comfort levels of these stakeholders in using telemedicine. At the meso level, costs for both healthcare organisations and patients, slow clinical acceptance, non-interoperable systems, lack of conformance to standards and faulty implementation strategies can impact whether telemedicine can be a practical option for care delivery.^30^ Finally, at the macro level, implementation challenges common in developing countries linked to infrastructural issues like unreliable internet connection in rural regions and lack of regulatory frameworks can impact the use and scaling of telemedicine services.

As healthcare proceeds through a transformative period at an unprecedented scale, our findings contribute to the growing literature to help professionals across the healthcare sector to understand factors that can influence the effective design, implementation and optimisation of ongoing telemedicine delivery efforts in India’s evolving technological landscape. We hope that our findings can better inform policy and practice regarding the role telemedicine can realistically play in the digital health transformation India aspires to.

## Data Availability

All data produced in the present study are available upon reasonable request to the authors

## Acknowledgements

The authors would like to thank the whole team at the Oxford India Centre for Sustainable Development, and particularly, Ms Vinita Govindarajan (Partnerships & Communications Manager), for their help with survey dissemination and valuable advice. Additionally, the authors would like to acknowledge: Dr Santanu Sen (former President of the Indian Medical Association); Dr Surekha Patil (Dean) of D.Y. Patil University School of Medicine and Dr Sriram Gopal (Head of Ob-Gyn Department); and Dr Vivek Bheemisetty (Orthopaedic surgeon at Zoi Hospitals, Hyderabad) for support with survey dissemination across India. The authors also thank Dr Vijay Patil (President of D.Y. Patil University) for supporting this study.

